# Model-based Cellular Kinetic Analysis of Chimeric Antigen Receptor-T Cells in Humans

**DOI:** 10.1101/2020.05.09.20096586

**Authors:** Can Liu, Vivaswath S. Ayyar, Xirong Zheng, Wenbo Chen, Songmao Zheng, Hardik Mody, Weirong Wang, Donald Heald, Aman P. Singh, Yanguang Cao

## Abstract

Chimeric antigen receptor (CAR)-T cell therapy has achieved considerable success in treating B-cell hematologic malignancies. However, the challenges of extending CAR-T therapy to other tumor types, particularly solid tumors, remain appreciable. There are substantial variabilities in CAR-T cellular kinetics across CAR-designs, CAR-T products, dosing regimens, patient responses, disease types, tumor burdens, and lymphodepletion conditions. As a “living drug”, CAR-T cellular kinetics typically exhibit four distinct phases: distribution, expansion, contraction, and persistence. The cellular kinetics of CAR-T may correlate with patient responses, but which factors determine CAR-T cellular kinetics remain poorly defined. Herein, we developed a cellular kinetic model to retrospectively characterize CAR-T kinetics in 217 patients from 7 trials and compared CAR-T kinetics across response status, patient populations, and tumor types. Based on our analysis results, CAR-T cells exhibited a significantly higher cell proliferation rate and capacity but a lower contraction rate in patients who responded to treatment. CAR-T cells proliferate to a higher degree in hematologic malignancies than in solid tumors. Within the assessed dose ranges (10^7^–10^9^ cells), CAR-T doses were weakly correlated with CAR-T cellular kinetics and patient response status. In conclusion, the developed CAR-T cellular kinetic model adequately characterized the multiphasic CAR-T cellular kinetics and supported systematic evaluations of the potential influencing factors, which can have significant implications for the development of more effective CAR-T therapies.

## INTRODUCTION

The success of immunotherapy, including chimeric antigen receptor (CAR)-T cell therapy, has fundamentally altered the landscape of cancer treatment and shows the potential to make previously incurable cancers now curable^1,2^. Chimeric antigen receptor (CAR), a genetically engineered receptor in T cells, consists of T-cell-activating domains (CD3ζ and costimulatory signals, such as CD28 or 4-1BB) and extracellular single-chain variable fragment (scFv) that binds to tumor-associated antigen^2,3,4^. The CAR equips the patient’s own T cells to recognize and subsequently eradicate cancer cells. While the mechanism of actions is not fully understood, evidence showed that CAR-T cells, once bound to the specific antigen on tumor cells, could release cytokines and other cell lysis mediators (e.g., granzyme B and perforin) that may directly kill antigen-expressing tumor cells. CD19-targeted CAR-T therapy has produced a high rate of durable remission in refractory B-cell malignancies, such as B cell acute lymphocytic leukemia (ALL) and chronic lymphocytic leukemia (CLL)^3,5^. The promising efficacy and the approval of two autologous CD19 CAR-T therapies (Yescarta and Kymriah) by the US FDA have motivated hundreds of follow-up CAR-T trials, including different CAR constructs, tumor antigen, and tumor types^2,6^. Among these CAR-T trials, there are about 73.8% (284/385) trials evaluating efficacy in hematologic malignancies and 26.2% (101/385) in solid tumors (clinicaltrials.gov, accessed Jan 2020).

Remarkable inter-subject and inter-trial variabilities in CAR-T cellular kinetics and therapeutic responses have been unfolded with the increasing number of CAR-T trials. For instance, CD19 CAR-T cell induced a significantly higher response rate in patients with ALL (> 80%) compared with CLL^2,3^. By contrast, CAR-T therapy has shown limited success in solid tumors, although several positive trials have been reported^7^. Although critical for the CAR-T clinical study design, the dose-response relationships of CAR-T therapy remain largely undefined^8-10^. Patients who received higher doses of CAR-T cells did not consistently see greater CAR-T cellular expansion and persistence, nor greater efficacy and long-lasting effect. Many other factors may confound the dose-response relationships, such as CAR-T constructs, lymphodepletion conditions, and baseline tumor burden. Some of these factors have been assessed in specific CAR-T trials, but it remains to be defined whether these factors could be generalized to other CAR-T therapies. To address these challenges, we performed a model-based analysis of CAR-T cellular kinetics and responses.

As a “living drug”, CAR-T cells rapidly proliferate upon antigen recognition, which distinguishes itself from traditional therapeutic modalities^11^. There are multiple phases in CAR-T cellular kinetics: distribution, expansion, contraction, and persistence. Each phase reflects an aspect of its functionality, as well as its interactions with host factors, such as tumor baselines and host immune functions. A rapid and high expansion of CAR-T cells is usually believed to be a positive sign for a response. The lymphodepleting therapy consisting of cyclophosphamide and fludarabine is routinely performed and sometimes relevant to CAR-T rapid cellular expansion^12–15^. The ensuing contraction phase after the expansion is related to mechanisms of activation-induced cell death (AICD) and the loss of antigen stimulation^16^. Durable responses are usually accompanied by long-term CAR-T cell persistence^10^. Along with the rapid clonal expansion, higher CAR-T cell exposures often elicit more adverse events, such as cytokine release syndrome (CRS) characterized by fever and multiple organ dysfunction and neurologic toxicity ^3,17^.

The traditional pharmacokinetic models are not applicable to characterize CAR-T cellular kinetics as it does not follow the typical disposition and elimination pathways as the conventional therapeutic molecules. An empirical cellular kinetics model has been developed by Stein et al^11^, which was further taken to evaluate the extrinsic and intrinsic factors that may impact the tisagenlecleucel expansion. Herein, we developed an empirical CAR-T cellular kinetics model based on two previously established immune dynamic models^11, 18^. We attempt to characterize the multi-phasic kinetics of CAR-T cells and systematically analyze the factors influencing CAR-T therapy in humans.

## METHODS

### Clinical data

Literature search covered CAR-T clinical trials with different targets and indications, including both hematologic and solid tumors. In total, seven clinical trials were included (Table 1) based on the following inclusion criteria: (1) Second generation CAR-T construct with one co-stimulatory domain; (2) Blood CAR-T abundance vs. time profiles were available; and (3) CAR transgene was quantitated by qPCR (copies/μg DNA). Pharmacokinetic data of CAR-T cells were digitized using WebPlot Digitizer (https://automeris.io/WebPlotDigitizer). The subject number, blood sampling intensity and duration, limit of quantification, and individual patient characteristics were summarized in Table S1. The quality of our digitization, in comparison with the original plots, was also evaluated in Figure S1.

**Table 1.** Overview of included CAR-T clinical trials. ALL, acute lymphocytic leukemia; BCMA, B-cell maturation antigen; CLL, chronic lymphocytic leukemia; CRS, cytokine release syndrome; DLBCL, diffuse large-B cell lymphoma; EGFR, epidermal growth factor receptor; EGFRvIII, epidermal growth factor receptor variant III; GBM, glioblastoma; MM, multiple myeloma; NSCLC, non-small-cell lung cancer; scFv, single-chain variable fragment.

| Sponsor | Construct | Indication | No. of Subjects | Dose | % Responders | CRS Toxicity | Clinical Trial & Reference |
| --- | --- | --- | --- | --- | --- | --- | --- |
| Novartis | Anti-CD19 scFv /4-1BB / CD3ζ | DLBCL | 86 | 0.1-6 x 10<sup>8</sup> | 52 | grade 3-4 (63%) | NCT02445248 (23) |
| University of Pennsylvania & Novartis | Anti-CD19 scFv /4-1BB / CD3ζ | CLL | 13 | 0.14-11 x 10<sup>8</sup> | 57 | grade 3-4 (43%) | NCT01029366 (10) |
| Novartis | Anti-CD19 scFv /4-1BB / CD3ζ | ALL (Pediatric) | 20 | 0.2-5.4 x 10<sup>6</sup>/kg;<br>0.03-2.6 x 10<sup>8</sup>/kg | NA | NA | NCT02435849<br>NCT02228096 (11) |
| Fred Hutchinson Cancer Research Center | Anti-CD19 scFv /4-1BB / CD3ζ | ALL (Adult) | 52 | 2 x 10<sup>5</sup>/kg;<br>2 x 10<sup>6</sup>/kg | 85 | grade 3-4 (19%) | NCT01865617 (14) |
| University of Pennsylvania & Novartis | Anti-BCMA scFv /4-1BB / CD3ζ | MM | 25 | 1) 1-5 x 10<sup>8</sup>;<br>2) 1-5 x 10<sup>7</sup>+Cy;<br>3) 1-5 x 10<sup>8</sup>+Cy | 1) 44;<br>2) 20;<br>3) 64 | grade 3-4 (32%) | NCT02546167 (15) |
| Chinese PLA General Hospital | Anti-EGFR scFv /4-1BB / CD3ζ | NSCLC | 11 | 0.45-1.1x10<sup>7</sup>/kg | 63.6 (PR/SD) | grade 3-4 (9.1%) | NCT01869166 (24) |
| University of Pennsylvania & Novartis | Anti-EGFRvIII scFv/4-1BB / CD3ζ | GBM | 10 | 1.75-5x10<sup>8</sup> | 33.3 | grade 3-4 (33.3%) | NCT02209376 (25) |

### Model structure

The model was built with a few adjustments of two previous models^11,18^. Model assumptions are: (1) Multiple phases of kinetics occur in chronological order; (2) Memory differentiation is not reversible during clinical trials. More specifically, circulating CAR-T cells first experience a transient decline due to tissue distribution after infusion (Fig. 1A).

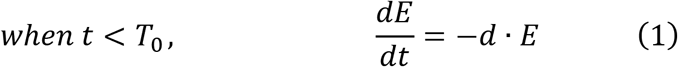

where *d* is distribution rate constant, T_0_ is distribution duration, and E is effector CAR-T concentration. The early distribution generally occurring immediately after administration only lasts for a couple of days and was not captured in most clinical trials. The early distribution phase was only depicted in the trial of DLBCL. In the model, for the trials without a clear early distribution phase, equation (1) was not included (T_0_ = 0).

After distribution, CAR-T cellular kinetics is governed by an exponential growth, up to the peak at time T_1_. It is assumed that there is negligible cell death and memory differentiation during the expansion phase (i.e., before T_1_).

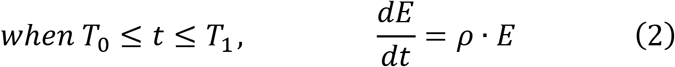

where ρ is proliferation rate constant, T_1_ is proliferation duration. To improve model estimation on the time constants, T0 and T_1_ were pre-determined based on each CAR-T profile.

**Figure 1.**
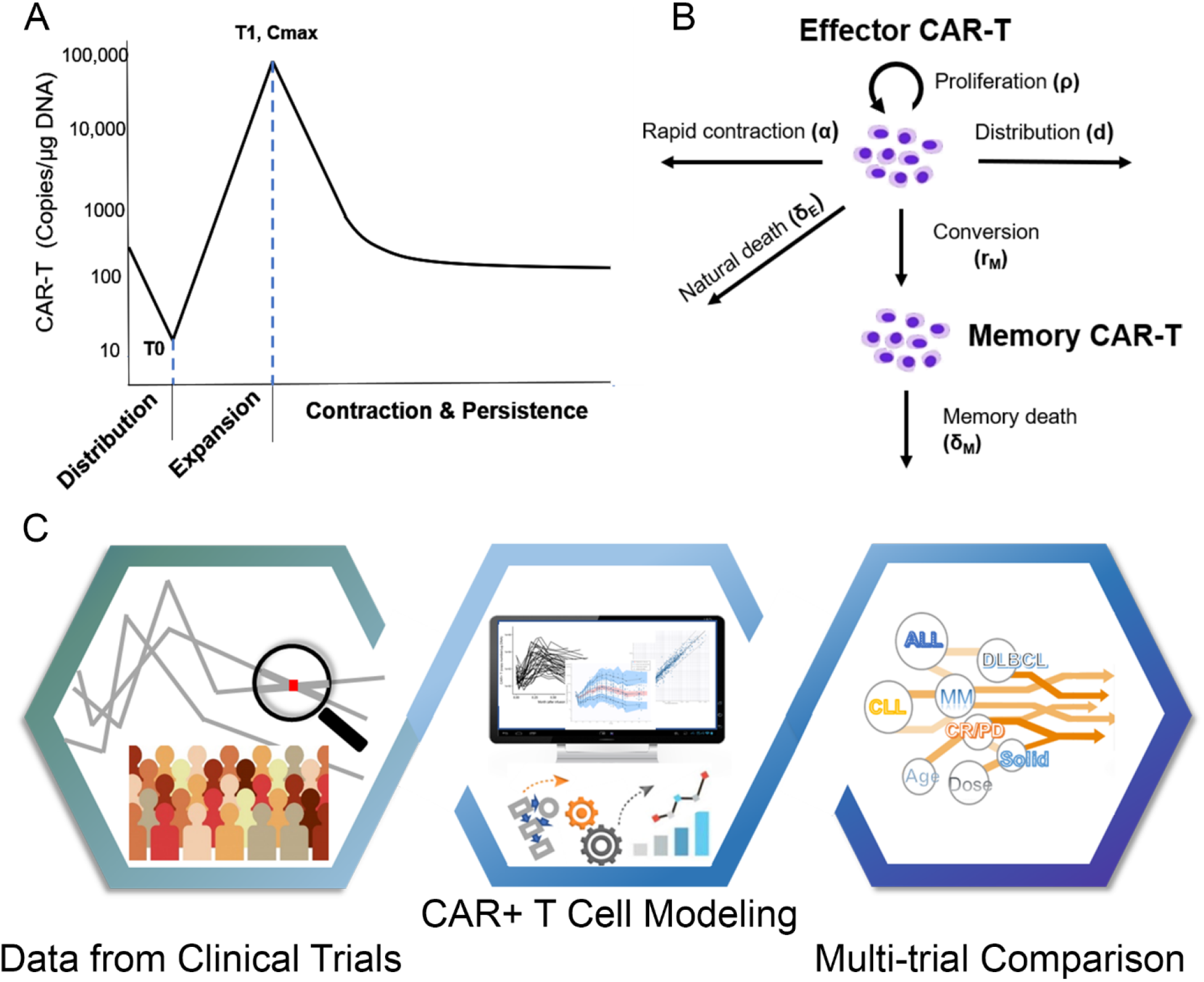
(A) A typical multi-phase kinetics of CAR-T cell: distribution, expansion, contraction and persistence phase; (B) Model structure for CAR-T kinetics; (C) Schematic diagram of three-step workflow for modeling and analysis.

In the phase of contraction and persistence, effector CAR-T cells are eliminated in three ways (Fig. 1B). AICD is a featured mechanism of the lymphocyte homeostasis after immune activation^19-21^, which mainly contributes to the initial rapid contraction (Fig. 1A). Meanwhile, a small fraction of effector CAR-T cells convert to memory cells or undergo a natural turnover.

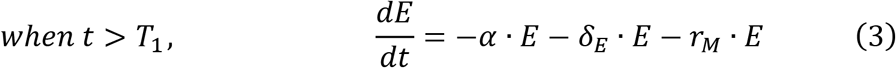

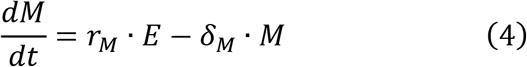

where α is rapid contraction rate constant, δ_E_ is the natural death rate constant of effector CAR-T, r_M_ is differentiation rate constant to the memory cell, M is memory-phenotypic CAR-T cell concentration, which indicates the CAR-T cells with longer persistence in the system. δ_M_ represents memory cell death rate constant. A biphasic contraction was observed in almost all individual CAR-T profiles, featuring a rapid contraction followed by a shallower differentiation phase (Fig. 1A). We did not assume that the rapid contraction stopped at the inflection point of the curves; therefore, α was allowed during the entire contraction. The terminal phase was associated with the death rate of the memory-phenotypic CAR-T cells.

### Data fitting and simulation

Distribution constant (d) was only applicable for the trial of DLBCL, and δ_E_ was fixed to 0.435 mon^-1^, according to a previous report^22^. Thus, there were in total of six parameters (d, C_max_, ρ, α, r_M_, and δ_M_) requiring estimated from the data. Random effects were also modeled on those parameters assuming log-normally distributed variance. A proportional residual error model was used with a log-normal distribution of the residuals. The model output was total CAR-T concentration (E + M). The initial concentration of the memory-phenotypic CAR-T cells M (0) was assumed to be zero, while the initial effector CAR-T concentration E (0) was set to:

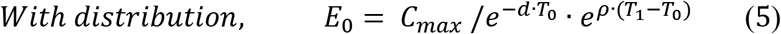

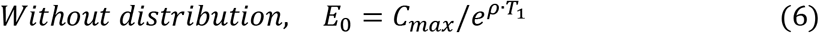

Population pharmacokinetic analyses were conducted using the stochastic approximation expectation maximization (SAEM) algorithm for nonlinear mixed-effect modeling, as implemented in Monolix version 2018R2 (Lixoft, France).

Parameter estimation was conducted separately by trial, and all estimates were summarized in Table S2. Model simulations were performed to illustrate the dynamics of the total, effector, and memory-phenotypic CAR-T for each CAR-T therapy and tumor type. The population values of parameters with and without inter-individual variability were applied to simulate concentration-time profiles of the total, effector, and memory-phenotypic CAR-T. Simulated CAR-T cellular kinetics between responders and non-responders were also compared. Monolix and R (3.5.3) were both used in simulation and graphing.

### CAR-T cellular kinetic variability

Because of large variabilities across CAR-T trials, data analysis was performed independently within each trial. We compared the CAR-T cellular kinetics between responders and non-responders and between hematologic and solid tumors, as well as across other potential factors that might contribute to inter-individual and inter-study variabilities. Six model parameters (C_max_, ρ, T_1_, α, r_M_, and δ_M_), which jointly determine the cellular profiles, were compared between groups with different responses. The definition of patient response status was summarized under Table S1. Patients were pooled into three categories of cancer types (ALL/CLL/MM, lymphoma, and solid tumors) to make a rough comparison of CAR-T proliferation and persistence across tumor types. Moreover, the correlations of model parameters with CAR-T cell dose, age, pre-treatment tumor burden, CD4: CD8 ratio in CAR-T products, and the lymphodepletion effect, were explored. Statistical analysis and data visualization were performed using R (3.5.3).

## RESULTS

### Clinical trial datasets and CAR-T cellular kinetics model

We digitized data with complete profiles of CAR-T cellular kinetics. In total, seven clinical trials with 217 patients were included in our final analysis, CAR-T therapies targeting hematological malignancies, lymphomas, and solid tumors^10,11,14,15,23-25^. The selected CAR-T therapies targeting different tumor antigens contained the same intracellular co-stimulatory domain 4-1BB belonging to the second generation of CAR-T construct (Table 1, Table S1, and Fig S1).

The multiphasic features in CAR-T cellular kinetics are similar, regardless of tumor types, dosing regimens, and sampling intensities. As shown in Figure 1A, the typical CAR-T cell kinetic profile comprises four distinct phases: early distribution, expansion, contraction, and persistence phases, which illustrate the physiological dynamics of CAR-T in the body upon injection: tissue distribution, proliferation, contraction, and memory-phenotypic differentiation, respectively^11^. The early distribution phase was characterized in the patients with DLBCL owing to intensive sampling in the early time points. In contrast, the expansion, contraction, and persistence phases were observed in all trials.

These features are well characterized by our CAR-T cellular kinetic model (Fig. 1B). The model structure originates from an immune-dynamic model reported previously with modifications for CAR-T cells^11, 18^. We fitted all individual data within each trial. Subsequently, we examined model-based parameter estimates with a series of *post-hoc* analyses on model estimates, patient responses, and many other potentially influencing factors. The whole workflow schema was summarized in Fig 1C.

### Model fitting and model parameters

Overall, the developed cellular kinetic model adequately characterized 217 individual CAR-T profiles. In the case of DLBCL, the model well-captured each of the phases, as shown in the fitting plots of representative patients (Fig 2A). The goodness-of-fit plot demonstrated good agreement between observed and model-predicted CAR-T concentrations (Fig 2B). Additionally, the visual predictive check (VPC) for the model showed good overall agreement between observed and simulated data in terms of percentiles of the individual distribution. The percentiles of observed data were close to the predicted percentiles and remained within the corresponding prediction intervals (Fig 2C). Individual fitting and model diagnostics plot for other trials were included in Figure S2-S7.

**Figure 2.**
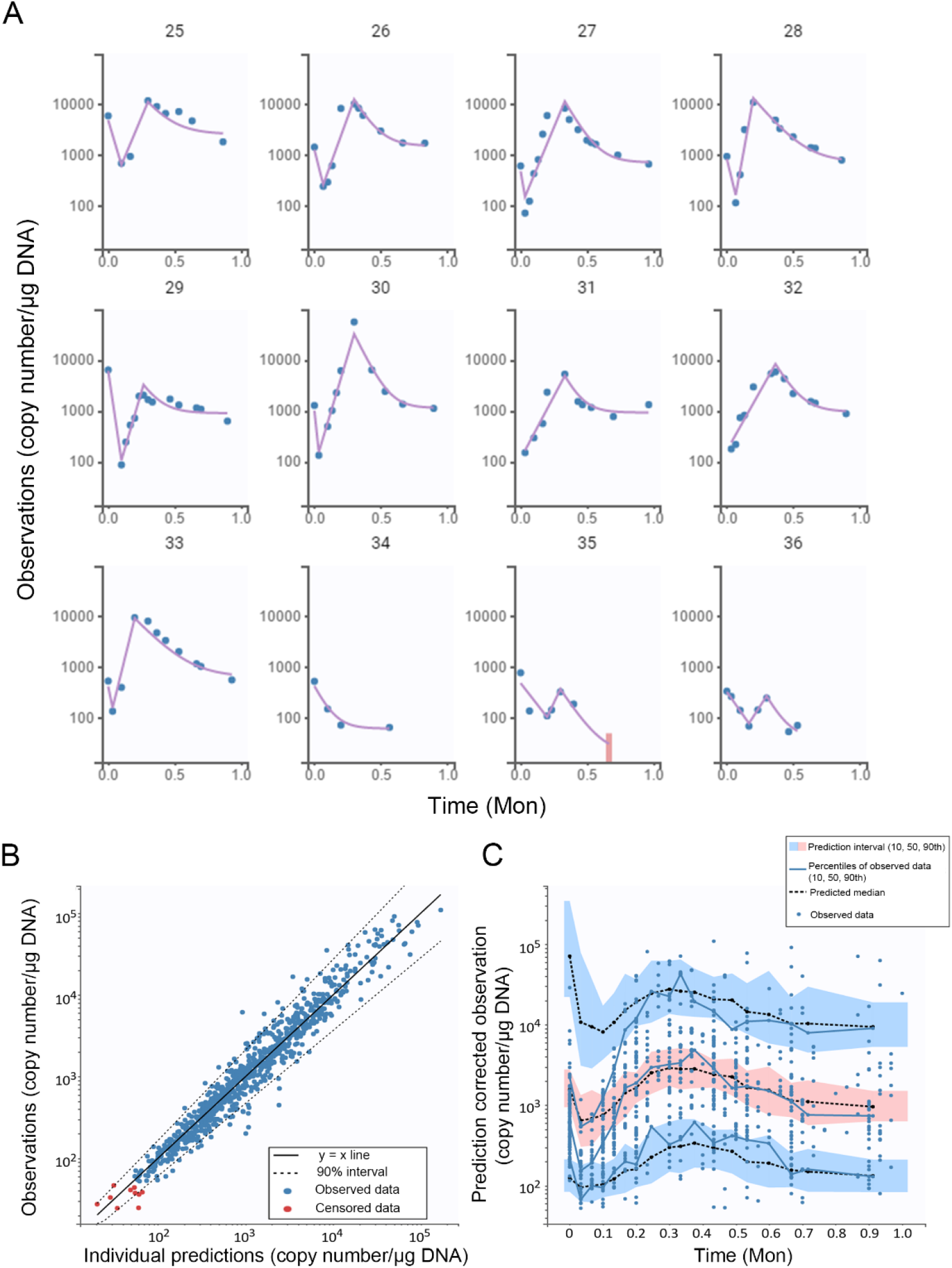
Model fitting and validation plots in the trial of DLBCL. (A) Individual fitting plots of representative patients; (B) The goodness-of-fit plot; (C) Visual predictive check (VPC) plot

Population parameter estimates, inter-individual variability (IIV), and estimation precision were listed in Table S2. Taking the MM patient data, for example, the population estimate of ρ is 17.0 mon^-1^ with an IIV of 0.354, and α is 4.74 mon^-1^ with an IIV of 0.863, while r_M_ and δ_M_ have relatively higher IIV, 1.68 and 1.26, respectively, mainly owing to the lack of a distinct persistence phase in many individuals. Overall, model parameters were reasonably estimated with acceptable precision and variability. The data in the contraction and differentiation phases were sparse in many patients, resulting in high errors of posterior individual parameters in these individuals, which was partially reflected in the VPC plots. Parameter estimates for all individual patients were shown in Figure 3.

**Figure 3.**
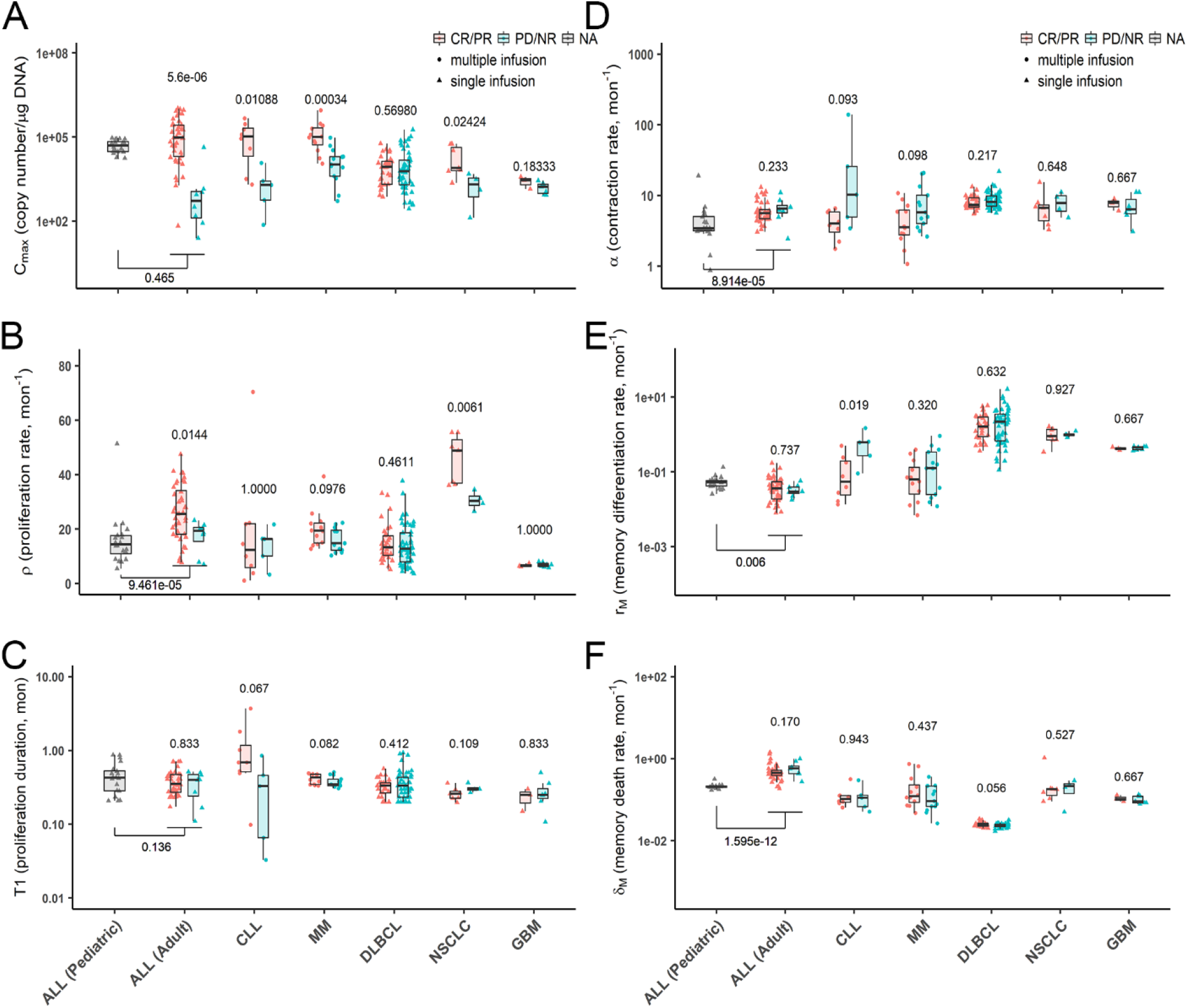
Individual model-estimated parameters for responders (CR/PR) and non-responders (PD/NR). Data from responders and non-responders are shown separately and compared for each trial. Two ALL trials were also compared, although response information was not available in the trial of ALL (Pediatric). Data distribution were described by boxplot (minimum, first quartile, median, third quartile, and maximum). p < 0.05 (Wilcoxon test) is considered as significant difference.

### Responders vs. Non-responders

Individual model parameters were compared by response status across trials to explore if responders (CR/PR) displayed different CAR-T cellular kinetics with non-responders (PD/NR) (Fig 3, and S8). For the distribution phase in DLBCL patients, responders had a significantly higher rate (d) of distribution with a shorter duration (T_0_) than those in non-responders (Fig S8). Of note, the samples at the very early time points may have contamination issues as some trials used the same tube to deliver the CAR-T cells and subsequently collect blood samples, so the CAR-T distribution rate should be interpreted with caution. During CAR-T expansion (Fig 3A-B), responders in many trials (4 out of 6) showed significantly higher proliferation capacities (C_max_). Namely, responders with ALL (adult), CLL, MM, and NSCLC had 159, 54.5, 9.59, and 3.57 folds increase of C_max_ than non-responders. Higher C_max_ in responders was partly due to higher proliferation rate (ρ), as in trials of ALL (adult), NSCLC, and MM, responders showed 1.33, 1.61 and 1.31 folds higher ρ, compared to non-responders. Responders with CLL or MM appeared to have 2.1 and 1.24 folds longer proliferation duration (T_1_), although they did not reach statistical significance, which partly explained a significantly higher C_max_ (Fig 3C). In addition to the higher proliferation, CAR-T cells in responders tended to have a relatively slower contraction rate (α). For example, the median α in responders of CLL and MM was only about 39.3% and 61.3% of those in non-responders, albeit no statistical significance (Fig 3D). The clinically relevant differences in CAR-T kinetic parameters should be evaluated in future studies. There were no statistically significant differences between responders and non-responders in terms of the memory cell differentiation (except for CLL) and death rate constants (r_M_ and δ_M_) (Fig 3E-F). The parameter estimates associated with the terminal phase (r_M_ and δ_M_) should be interpreted with caution due to the limited sampling durations in most of the CAR-T trials.

### Hematologic malignancies *vs*. Solid tumors

In hematologic malignancies (ALL/CLL/MM), CAR-T cells seemed to proliferate for a longer duration (T_1_) and reach a significantly higher proliferation capacity (C_max_) than in lymphoma and solid tumors (Fig 4A and C). Hematologic malignancies appeared to be associated with a higher CAR-T proliferation rate (ρ) than lymphoma (1.5 folds), but not solid tumors partly due to considerable variability of CAR-T proliferation rate in NSCLC and GBM (Fig 4B). By contrast, solid tumors were associated with a higher CAR-T contraction rate (α) by 1.3 folds (Fig 4D). These comparisons were not intended to inform CAR-T properties nor product qualities as these CAR-T trials had different targets, CAR-T designs, manufacturing processes, and patient characteristics.

**Figure 4.**
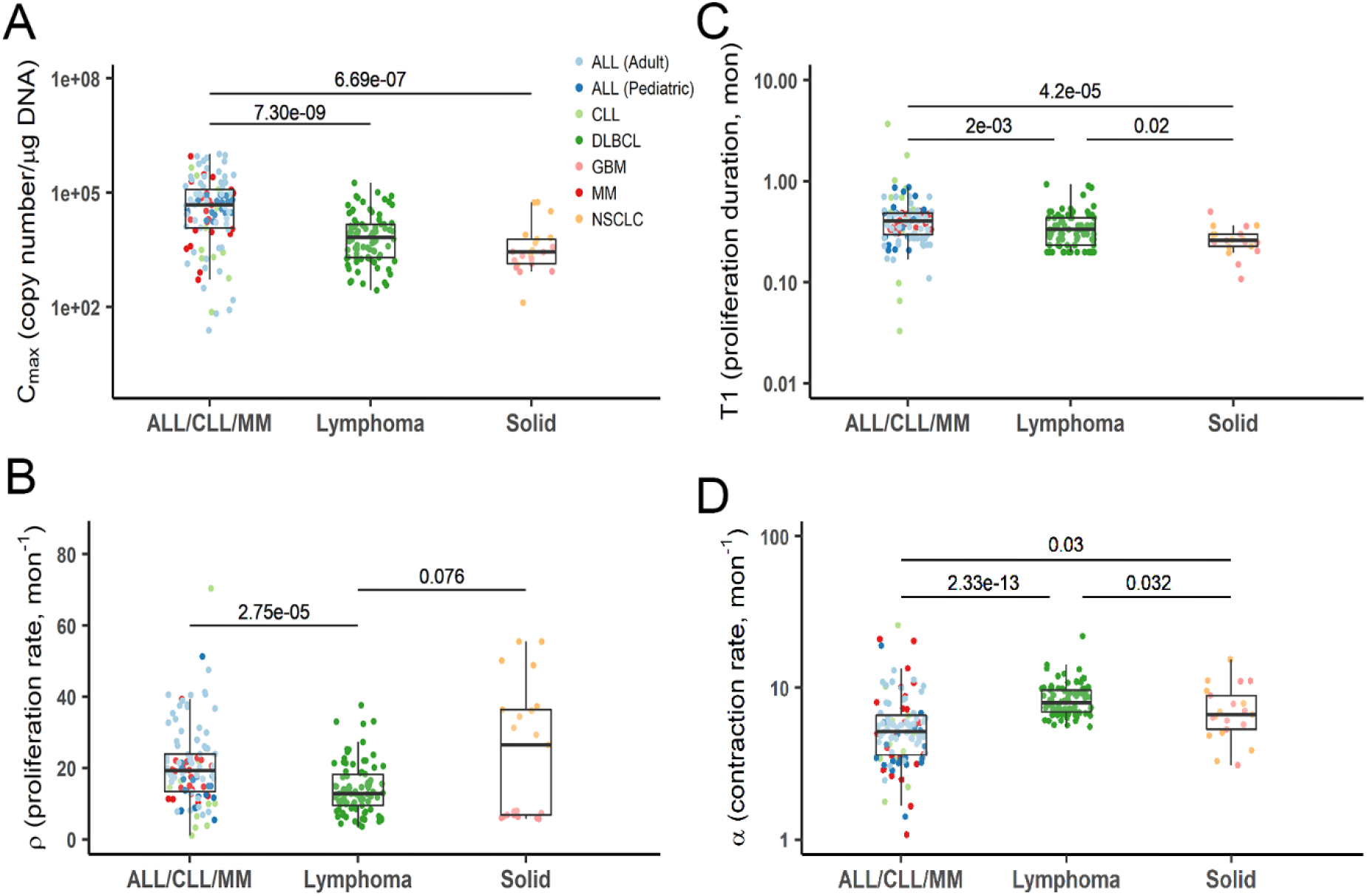
Individual model-estimated parameters of three tumor types (ALL/CLL/MM, lymphoma and solid tumors). Data from corresponding trials were pooled into three main categories. Data distribution were described by boxplot (minimum, first quartile, median, third quartile, and maximum). p < 0.05 (Kruskal–Wallis test) is considered a significant difference.

### Effect of CAR-T doses, baseline tumor burdens, and CD4: CD8 ratios

The influence of CAR-T cell doses on the model parameters was assessed in four trials (CLL, MM, GBM, and NSCLC) with individual dosing records. Due to high variabilities, CAR-T doses in the range of 10^7^ – 10^9^ cells/patient were weakly correlated with patient responses, regardless of tumor types (Fig 5A). Furthermore, neither any parameter showed significant dose correlation within each trial (Fig S9).

**Figure 5.**
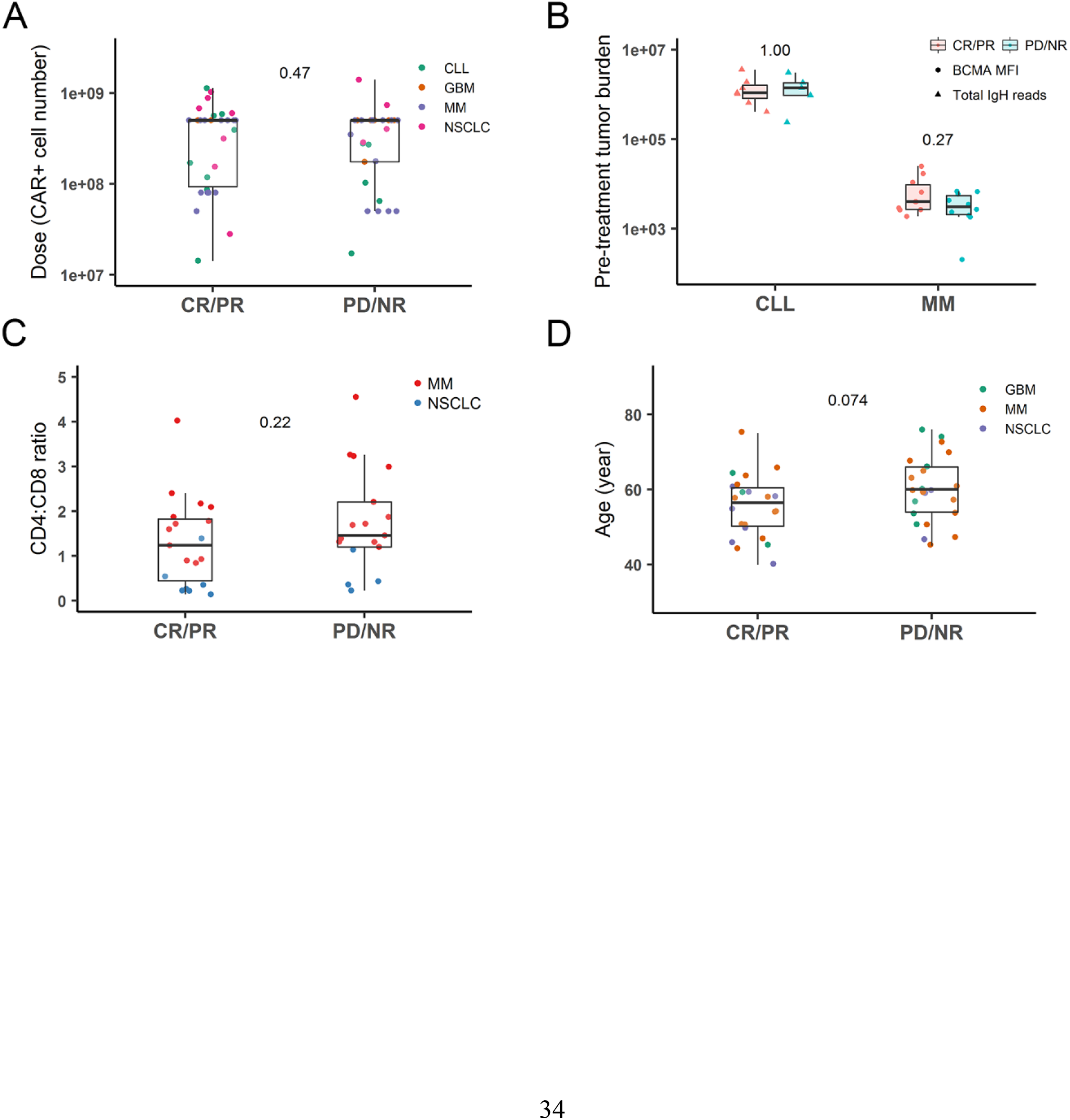
Patient covariates for responders (CR/PR) and non-responders (PD/NR). (A) A comparison for individual dosing by the response in the trials of CLL, GBM, MM, and NSCLC; (B) A comparison for pre-treatment tumor burden by the response in the trials of CLL and MM. Tumor burden values were not comparable across trials, as they were measured differently, indicated as “BCMA MFI” and “total IgH reads”, respectively; (C) CD4: CD8 ratio was compared by the response in the trials of MM and NSCLC; (D) Patient age was compared by the response in the trials of MM, GBM and NSCLC.

Pre-treatment tumor burdens were individually measured in the CAR-T trials of MM and CLL. Similar as TCR-mediated T cell activation, the size of tumors is generally considered as a crucial factor for efficient CAR-T cell activation and expansion. Patients with substantial tumor burden frequently suffered from high cytokine release syndromes in several trials, suggesting rapid and significant CAR-T expansions in these patients^26-28^. However, based on the data utilized within our analysis, in MM and CLL patients, pre-treatment tumor burdens were not statistically correlated with the potential of CAR-T proliferation (Fig S10). For CLL, patients with a higher tumor burden showed an uptrend toward a higher C_max_, but no significance was reached (Fig S10B). There was no apparent difference in pre-treatment tumor burdens between responders and non-responders either (Fig 5B). The reasons for this inconsistency were not entirely clear, which was probably associated with the uses of circulating tumor antigens as biomarkers for tumor burden quantification in the analyzed studies.

The CD4: CD8 ratios in the infused CAR-T cell products were also quantified in both MM and NSCLC trials. Responders appeared to have CD4: CD8 rates closer to 1 while non-responders (Fig 5C) showed relatively higher and diverse ratios, although no statistical difference was detected. Similarly, the CAR-T proliferation capacity (C_max_) and rate constant (ρ) tended to correlate with CD4: CD8 ratios negatively, but significance was not reached in either trial (Fig S11A-B).

### Other factors (patient age and lymphodepletion)

The effect of other factors, such as patient age and lymphodepletion treatment, were evaluated similarly. To some extent, patient age is associated with immune functions^29^. Compared with adult patients with ALL, pediatric patients with ALL displayed a significantly lower proliferation rate, but a lower contraction, a higher memory differentiation, and lower memory death rate (Fig 3). By contrast, for adult patients between 40 and 80 years, age did not seem to correlate with any kinetic parameters in any CAR-T trial (Fig S12), which is consistent with previous findings^30^. Interestingly, patients in the CR/PR group appeared to be younger than patients in the PD/NR group (Fig 5D), albeit no statistical significance.

Lymphodepletion has become a standard procedure prior to CAR-T cell infusion to minimize immune-mediated CAR-T cell rejection^31^. In the MM trial, lymphodepletion effect was evaluated by comparing two cohorts of patients with the same CAR-T doses with or without lymphodepletion. The data showed that lymphodepletion resulted in a higher CAR-T proliferation capacity (C_max_) in MM patients (Fig S13A). However, similar results were not observed in the NSCLC trial because lymphodepletion was only applied to NSCLC patients with larger tumor sizes. About the lymphodepletion regimen, fludarabine, in addition to cyclophosphamide, tended to enhance CAR-T proliferation (Fig S14).

### Parameter correlations

Statistical correlations among model parameters were explored (Fig S15). As expected, higher CAR-T proliferation capacity (C_max_) were associated with faster proliferation rates (ρ) and longer proliferation durations (T_1_), considering that C_max_ is a function of ρ and T_1_ (Fig S15A-B). Proliferation durations (T_1_) were negatively correlated with proliferation rate constant (ρ) (Fig S15C), resulting in a comparable proliferation potential across individual, and suggesting a host-restricted proliferation capacity in humans. CAR-T contraction rate constant (α) tended to mildly decrease with an increased ρ and C_max_ (Fig S15D and E). In addition, the memory differentiation rate constant (r_M_) negatively correlated with the memory cell death rate constant (δ_M_), contraction rate constant (α), as well as C_max_ in most trials (Fig S15F-H).

### Model simulation

The inter-individual and inter-study variability were further explored and visualized by simulating 1000 virtual patients for each trial (Fig S16). The simulation was performed using the population-typical values of each trial’s model parameters, along with the estimated inter-individual variabilities (IIV). The distribution profiles of actual populations were located within 10^th^ – 90^th^ percentile intervals.

To distinguish the kinetics of effector CAR-T and memory-phenotypic CAR-T cells, the profiles of total CAR-T cells and those two subpopulations were simulated for each trial (Fig 6). Although the terminal phase was not entirely followed in some trials, the simulations still help to characterize the rapid contraction of effector CAR-T and the persistence of memory-phenotypic CAR-T cells within, as well as beyond the study duration of clinical trials.

**Figure 6.**
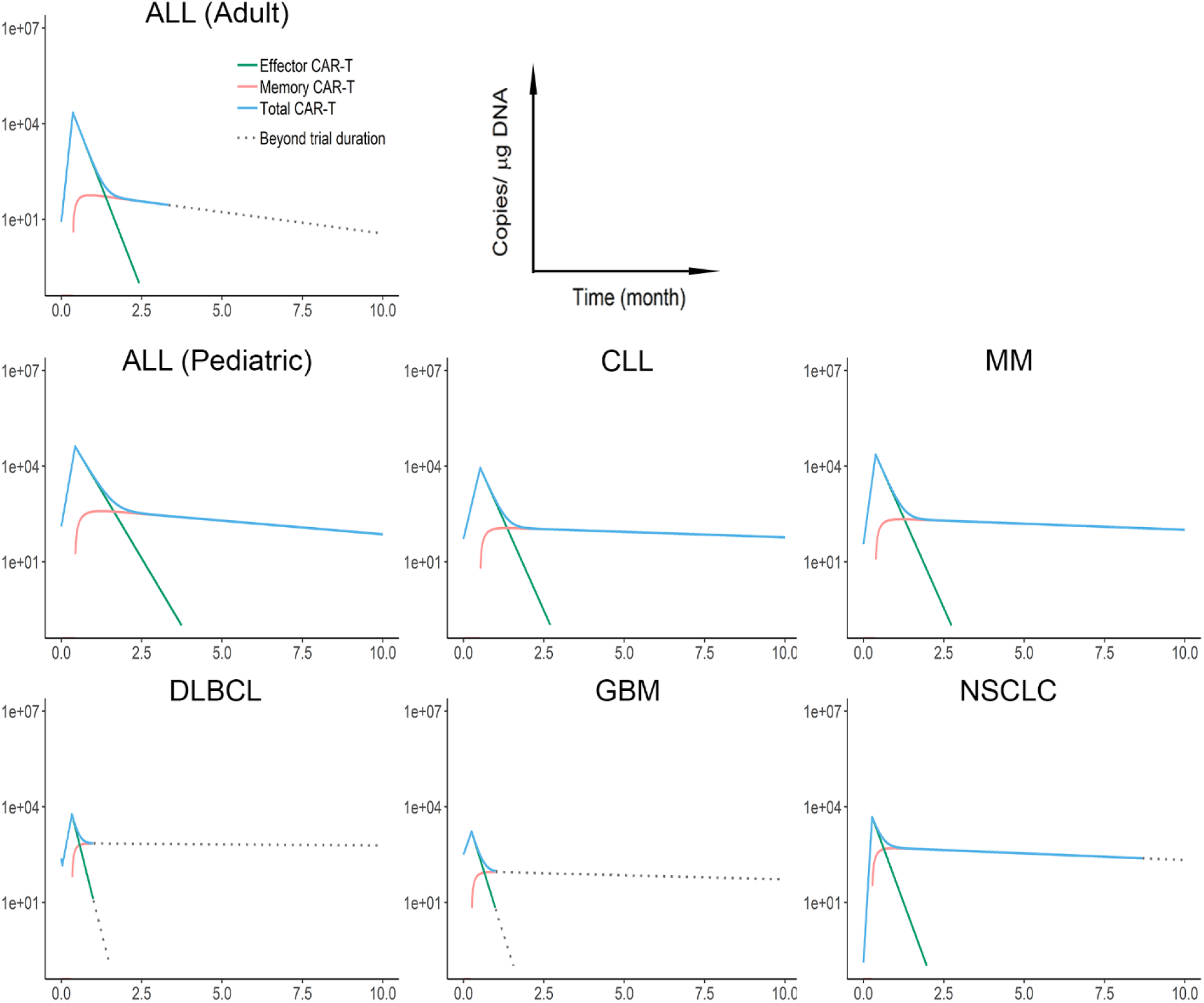
The simulated kinetics of effector CAR-T (green), memory-phenotypic CAR-T (red), and total CAR-T (blue) for each clinical trial. Simulations were performed based on the population typical values of parameters. CAR-T cell kinetics were simulated until 10 months. Dotted lines represent the duration beyond the period of the clinical trials.

Compared with adult ALL patients, pediatric ALL patients seemed to have a slower contraction but higher memory differentiation rates (Table S2, Fig 3 and 6). A relatively higher CAR-T proliferation potential was observed in ALL patients than in CLL patients (Table S2, Fig 3 and Fig 6). This is consistent with the observation that CD19 CAR-T therapy exhibited a higher response rate in ALL than in CLL. The interpretation of extrapolated profiles of CAR-T persistence in DLBCL and GBM patients must be performed with caution due to the short duration of sampling. Furthermore, CAR-T kinetics of responders and non-responders were simulated and compared (Fig S17A). Responders in the trial of ALL and MM displayed a higher peak of effector CAR-T, a higher formation of memory CAR-T, and a slower contraction rate of effector CAR-T. As considerable heterogeneity of effector and memory CAR-T kinetics was observed across trials (Fig 6), the inter-individual heterogeneity of those populations was simulated (Fig S17B).

## DISCUSSION

CAR-T therapy has achieved exceptional success in lymphoid B cell malignancies, but numerous challenges exist in extending this therapy to other tumor types, particularly solid tumors^32^. There exist substantial variabilities in CAR-T cellular kinetics and patient responses. Some experiences have been gained during clinical practice. For example, compared to the first-generation, second-generation CAR consisting of an intracellular costimulatory domain (either CD28 or 4-1BB) exhibited much higher CAR-T cell expansion and persistence, which appeared to be the drive for the improved response probability and efficacy^3,33^. Lymphodepletion prior to CAR-T infusion has been routinely applied to eliminate the native lymphocyte competition and facilitate CAR-T cell rapid expansion, which often results in enhanced patient response^13-15^. However, the key determinants governing CAR-T cellular kinetics and patient responses remain to be characterized, particularly considering which showed high variabilities and were confounded by a broad range of factors. Moreover, the quantitative impact of these factors on CAR-T cellular kinetics as well as the clinical outcomes has not been systematically analyzed yet, which is sometimes inconsistent across CAR-T types, dosing regimens, and pre-treatment tumor burdens.

It is difficult to perform a direct comparison of clinical outcomes across multiple trials with distinct CAR-T cell products, dosing regimens, tumor types, and heterogeneous patient populations. In this report, we developed a cellular kinetic model to characterize the pharmacokinetic behaviors of CAR-T cells, and after that, quantitatively and systematically evaluated CAR-T cellular kinetics and clinical outcomes across patient response statuses (Fig 1B and C). Our work offered a means to systematically characterize CAR-T kinetics-response relationships as well as the potential influencing factors across multiple CAR-T trials.

Unlike other therapeutic modalities, CAR-T cells that are capable of proliferation and differentiation in the system typically exhibit multiphasic kinetics. Moreover, CAR-T proliferation potential and systemic exposure are not strictly dose-dependent. Conventional compartmental pharmacokinetic models are therefore not strictly applicable to characterize CAR-T cellular kinetics. The developed model can recapitulate the multiple functional phases of CAR-T cells before, during, and after encountering tumor antigens. The model predicts the phases of distribution, expansion, and biphasic contraction, which consists of rapid contraction and memory cell differentiation and a patient-specific duration of each phase (Fig 1A). Although cell proliferation may take place during other phases or memory-phenotypic cells may be formed during the early phase, our model set T_1_ to separate proliferation phase and contraction phase to keep the model semi-mechanistic, yet simple, which enabled the model to capture the diverse CAR-T profiles (Fig 2 and S2-S7).

Patient response status was closely associated with CAR-T cellular kinetics (Fig 3 and S8). Responders were associated with an average of higher CAR-T expansion capacity and higher proliferation rate constant than non-responders (Fig 3A and B). A relatively lower contraction rate constant was observed in responders (Fig 3D). Although the early distribution phase was attainable only in the DLBCL CAR-T trial, the responders exhibited significantly faster distribution than non-responders (Fig S8). The mechanism for this observation remains unknown. More intensive samplings right after CAR-T infusion is preferable to confirm and fully capture the rapid distribution phase in future CAR-T trials. Of note, there were high inter-study variabilities in model parameters (Fig 3 and Table S2), particularly for r_M_ and δ_M_, which were estimated with high biases in CAR-T trials owing to the sparse sampling during the terminal phase.

Hematological cancers exhibited a higher CAR-T proliferation capacity (C_max_) and a lower contraction rate (α) compared with that in solid tumors (Fig 3, 4A and D), which may be attributed to the fluidic form of hematological cancers that enable the fast and high antigen accessibility and presentations upon CAR-T injection. In solid tumors, cancer cells are often spatially restricted^34^, providing poor antigen accessibility to CAR-T cells. CAR-T cells have to extravasate into the tumor bed, migrate toward the tumor cells, contact, and then engage the tumor cells^7,35^. Differences in tissue accessibility and delivery rate for CAR-T cells may account for large variability on proliferation rate constant (ρ) between two solid tumor trials (Fig 4B). Cell proliferation duration (T_1_) was the most conservative parameter across CAR-T trials and patient populations (Fig 3C), while blood cancers experienced more extended proliferation than lymphoma and solid tumors (Fig 4C).

Substantial inter-individual variabilities in CAR cellular kinetics were observed (Fig 3 and S16). The sources of variability and the influencing factors were analyzed. The CD4: CD8 ratios and lymphodepletion conditions showed a certain degree of influence on CAR-T expansion and differentiation, while pre-treatment tumor burden did not (Fig S10, S11, S13, and S14). When comparing multiple trial results in the dose range of 10^7^ ~ 10^9^, neither CAR-T cell expansion nor contraction was associated with CAR-T doses (Fig S9). Although a steep dose-response relationship was speculated^2^, a solid dose-dependency has not yet been found in many previous analyses^30,36^, potentially a result of variability in the T cell subset composition. Nevertheless, in the trial of MM^15^, cohorts 2 and 3 were treated with cyclophosphamide before the injection of a low (1-5 × 10^7^) or high dose (1-5 × 10^8^) of CAR-T cells. The high-dose cohort exhibited a significantly higher proliferation rate (ρ), and higher capacity (C_max_) compared to the low-dose cohort (data not shown). Studies also observed limited CAR-T expansion at a low dose (< 5 × 10^7^)^37, 38^, and no CAR-T expansion at very low doses (< 10^6^)^39^, indicating a potential threshold of dose to assure rapid CAR-T expansion and a steep dose-response curve for CAR-T therapy^2,35,40^. Similarly, although patient age was not correlated with any parameter in adult patients (40 - 80 years old), pediatric patients showed significantly lower CAR-T contraction than adult patients (Fig 3 and S12), which is probably due to the immature pediatric immune system that creates a more friendly environment to the persistence of CAR-T cells.

Our analysis has two significant limitations. First, the unbalanced sample size and blood sampling intensity may result in bias in the statistical comparisons in multiple trials. We thus drew one of our primary conclusions across patient response status within each trial (Fig 3) and made the comparisons stratified by trials (Fig 4 and 5). Besides, we performed a simulation study to make sure the relatively fewer samples in some trials did not influence the model fittings and parameter estimates (Fig S18). Second, the developed model was still mostly empirical. We considered two subpopulations of CAR-T cells in the model, as did in the previous model^18^. However, the biological distinctions of effector and memory populations remain unclear, so we defined the memory subpopulation as memory-phenotypic cells to reflect the CAR-T cells with extended persistence. The model included the early distribution phase and the natural death rate of effector cells, making it different from the previously developed models^11,18^. The contraction and persistent phases are mathematically exchangeable but with different parameter interpretation in comparison with previous models. Along with the rapid expansion of CAR-T candidates, more mechanistic cellular kinetics and dynamics models are warranted to disentangle the complicated relationships between target engagement, tumor burdens, as well as the dynamic native immune systems^35^.

In conclusion, the developed cellular kinetic model adequately characterized the CAR-T profiles in humans. The potential sources of variabilities in CAR-T cellular kinetics were systematically analyzed across patient populations and tumor types. Our analysis provided a starting point to understand further CAR-T cellular kinetics and kinetic-response relationships, which have implications for future CAR-T development and clinical optimization.

## Study Highlights

### • What is the current knowledge on the topic?

Despite the initial success of CAR-T therapy in B-cell malignancies, CAR-T cellular kinetics and the determining factors on CAR-T cellular kinetics and patient response remain poorly defined. As a “living drug”, traditional pharmacokinetic models do not apply to the analysis of CAR-T cellular kinetics.

### • What question did this study address?

This study aimed to develop a cellular kinetic model to characterize the multi-phasic kinetics of CAR-T cells and the relationship between CAR-T cellular kinetics and patient responses, and investigate the factors affecting CAR-T cellular therapies across patient populations and tumor types.

### • What does this study add to our knowledge?

Our results showed that patients who are responding exhibit a higher CAR-T expansion capacity and a greater proliferation rate, and a lower contraction rate than non-responders. CAR-T cells proliferate at a relatively higher rate in hematologic malignancies than in solid tumors.

### • How might this change clinical pharmacology or translational science?

Patient response to CAR-T therapies was closely associated with CAR-T cellular proliferation and persistence. The influencing factors identified to CAR-T cellular kinetic should have substantial implications for the development and clinical use of CAR-T therapies.

## Author Contributions

C.Y., SAP., H.D., W.W., L.C., A.V. S., Z.X., C.W., Z.S., and M.H. wrote the manuscript; C.Y, S.A.P., H.D., W.W., and L.C. designed the research; L.C. and C.Y. performed the research; L.C. and C.Y analyzed the data.

## Data Availability

All the data analyzed in this manuscript was from the published literature, which were all cited in the manuscript.

## Acknowledgement

We acknowledged all the patients, families, physicians, and medical staff that contributed to these trials.

